# Improving discriminative ability in mammographic microcalcification classification using deep learning: a novel double transfer learning approach validated with an explainable artificial intelligence technique

**DOI:** 10.1101/2025.08.05.25332967

**Authors:** K. Arlan, M. Björnström, T. Mäkelä, T. J. Meretoja, K. Hukkinen

## Abstract

**Background:** Breast microcalcification diagnostics are challenging due to their subtle presentation, overlapping with benign findings, and high inter-reader variability, often leading to unnecessary biopsies. While deep learning (DL) models - particularly deep convolutional neural networks (DCNNs) - have shown potential to improve diagnostic accuracy, their clinical application remains limited by the need for large annotated datasets and the “black box” nature of their decision-making.

**Purpose:** To develop and validate a deep learning model (DCNN) using a double transfer learning (d-TL) strategy for classifying suspected mammographic microcalcifications, with explainable AI (XAI) techniques to support model interpretability.

**Material and methods:** A retrospective dataset of 396 annotated regions of interest (ROIs) from full-field digital mammography (FFDM) images of 194 patients who underwent stereotactic vacuum-assisted biopsy at the Women’s Hospital radiological department, Helsinki University Hospital, was collected. The dataset was randomly split into training and test sets (24% test set, balanced for benign and malignant cases). A ResNeXt-based DCNN was developed using a d-TL approach: first pretrained on ImageNet, then adapted using an intermediate mammography dataset before fine-tuning on the target microcalcification data. Saliency maps were generated using Gradient-weighted Class Activation Mapping (Grad-CAM) to evaluate the visual relevance of model predictions. Diagnostic performance was compared to a radiologist’s BI-RADS-based assessment, using final histopathology as the reference standard.

**Results:** The ensemble DCNN achieved an area under the ROC curve (AUC) of 0.76, with 65% sensitivity, 83% specificity, 79% positive predictive value (PPV), and 70% accuracy. The radiologist achieved an AUC of 0.65 with 100% sensitivity but lower specificity (30%) and PPV (59%). Grad-CAM visualizations showed consistent activation of the correct ROIs, even in misclassified cases where confidence scores fell below the threshold.

**Conclusion:** The DCNN model utilizing d-TL achieved performance comparable to radiologists, with higher specificity and PPV than BI-RADS. The approach addresses data limitation issues and may help reduce additional imaging and unnecessary biopsies.

## Introduction

Microcalcifications are tiny, variable, often nonspecific yet critically important signs of both benign and malignant changes of the breast. Up to 75% of occult early breast cancer changes, particularly ductal carcinoma in situ (DCIS), present solely as microcalcification on mammograms (1). Breast Imaging-Reporting and Data System (BI-RADS) is a widely used tool that aids radiologists standardize the reporting of breast image findings, including microcalcifications among others. For suspicious microcalcifications BI-RADS categories 4a, 4b, 4c and 5 are used. Category 4a lesions have a 3–10% likelihood of malignancy, category 4b 11–50%, category 4c 51–94%, and category 5 greater than 95%. To enhance visibility, additional imaging techniques, such as a true lateral view, spot compression with fine focus magnification (spot), or tomosynthesis, are also often recommended. However, multifaceted pathophysiology of DCIS and the limitations of BI-RADS lexicon result in low specificity, with positive predictive values (PPV) ranging from 20 to 40% and substantial inter-reader variability (2,3). Consequently, stereotactic vacuum-assisted biopsy (SVAB) is commonly performed as the gold standard diagnostic procedure of microcalcifications. However, SVAB is a costly, time-consuming, complication-prone, and anxiety causing procedure.

Advances in computer image analysis, increased computational power, and the availability of large image datasets have significantly enhanced the performance of deep learning (DL) methods, particularly deep convolutional neural networks (DCNN), across numerous medical fields (4). Consequently, several DCNN have shown promising results in classifying suspicious microcalcifications with improved specificity and PPVs (5–8).

Training DCNN from scratch to perform efficiently on specific tasks (e.g. detection and characterization of suspected microcalcifications on mammogram) requires large amounts of labeled images, complex training procedures, heavy computational power, considerable time and budget (5). Moreover, the creation, access, storage, use and maintenance of medical image databases are strictly regulated by personal data protection and intellectual property laws (9). As a result, the limited access to specific training datasets may lead to overfitting, reducing the accuracy of the models.

Transfer learning (TL) is a technique that allows to reuse learned knowledge of pre-trained DCNN for specific tasks. The method employs a DCNN model pre-trained on a large, generic image dataset (i.e. ImageNet) and fine-tunes using task-specific training data (i.e benign and malignant microcalcification cases) to enable its application to a specific task (i.e. labeling suspicious microcalcifications). TL enhances model accuracy, particularly if trained on a limited amount of data (10,11). Furthermore, recent studies suggest that TL is more effective when the source and target datasets are similar (12,13). Thus, instead of fine-tuning a pre-trained DCNN model directly with a limited amount of training data (i.e. labeled microcalcification cases), an intermediate step of training on a more specific image dataset (i.e. labeled mammogram database) can further enhance its performance. This so-called “double transfer learning” approach may improve the DCNN’s performance for tasks with limited amount of training data.

Neural networks consist of many layers connected to each other with lots of nonlinear intertwined connections. As a result, it’s unfeasible to fully comprehend how the system comes to its conclusion (14). Therefore, DL is often considered as a “black box”. Uninterpreted and unexplained DL models may be biased, that go unnoticed which is unacceptable in high-stakes fields such as medicine. Gradient-weighted Class Activation Mapping (Grad-CAM) is a technique that provides visual explanations for DCNN decisions, making them more transparent (15). The field of explainable artificial intelligence (XAI) is growing due to the need to explain DL predictions in a form understandable to humans and integrate it into clinical practice (16,17).

This study aimed to develop a DL-based network utilizing the double transfer learning concept to predict malignant microcalcifications in the BI-RADS 4 subset. We compared the performance of DL with that of radiologist and analyzed Grad-CAM maps to confirm the validity of the new model.

## Materials and methods

This study did not require an ethics committee permission due to its retrospective nature but was reviewed and approved by Research Chair of the Helsinki University Hospital. The analysis was performed in accordance with the guidelines of the local institutional review board and the principles expressed in the Declaration of Helsinki.

### Database creation

The local image database was constructed from mammograms of the patients with suspected microcalcifications, initially identified through screening, clinical (primary care and private), or follow-up mammography after breast cancer treatment and that were referred to Women’s Hospital radiological department of Helsinki University Hospital (HUH) between 2016 and 2019 for SVAB procedure. Other mammographic abnormalities related to microcalcifications such as masses or architectural distortions were excluded from the study.

Image data was retrieved from Helsinki University Hospital Picture Archiving and Communication System (PACS). We used cranio-caudal (CC), medio-lateral oblique (MLO), and true lateral (LM) full-field digital mammography images to construct local labelled database. Spot images were not included. The mammograms were acquired from breast imaging systems from four different vendors: Hologic, Planmed, GE Healthcare, and Siemens.

Highly experienced breast radiologist selected benign and malignant cases for database creation. A radiology resident then reviewed all microcalcification cases eligible for the study, cropped each lesion by applying a square region of interest (ROI) to the area of suspected microcalcifications and annotated each cropped image with binary classification based on histopathology (benign = 0, malignant = 1). Another breast radiologist, who was not involved in dataset preparation, and was blinded to final histopathology, reviewed all test cases and categorized microcalcifications using the BI-RADS lexicon (categories 3-5)

The final diverse dataset comprised 396 segmented images from 194 patients. To develop and evaluate the performance of DL model, the dataset was randomly split into training and test datasets (24% for the test dataset with 50/50 proportion of benign and malignant cases).

### Development of DL model and neural network training

In this study, we developed and validated a DCNN for the classification of suspected mammographic microcalcifications using a novel d-TL strategy. This approach combines pretrained knowledge from both natural image domains and medical imaging datasets, enabling robust performance despite the limited size of domain-specific data. A detailed description of the architecture and training procedure will be provided in a separate methodological manuscript (in preparation).

### Performance metrics

For the test set, we used final pathology as the ground truth and calculated the sensitivity, specificity, accuracy, negative predictive value (NPV), and positive predictive value (PPV) with 95% confidence intervals (CI) for both the DL model and radiologist. We calculated ROC curves by varying the network’s confidence threshold.

### XAI technique

Saliency maps were generated using the Grad-CAM, a well-established visualization-based XAI method (15). Grad-CAM computes the regions within the input image that yield the highest impact on the output layer of the neural network (18). This process produces a heatmap that assigns a score to each input pixel according to its relevance in the decision-making process (19). False negative and false positive cases of the DL model were reviewed using Grad-CAM heatmaps. For the visual analysis, a probability threshold of 0.76 was selected for the activation maps, which corresponded to the optimal threshold of ROC curves.

## Results

The diagnostic performance of DCNN compared to radiologist is summarized in Tables 1 and 2.

**Table 1.** Diagnostic performance of DCNN model and radiologist for classifying suspicious microcalcifications (Test dataset N = 47).

|  | Sensitivity (%) | Specificity (%) | PPV (%) | NPV (%) | Accuracy (%) |
| --- | --- | --- | --- | --- | --- |
| DCNN model | 65 (42-84) | 83 (63-95) | 79 (59-91) | 71 (58-82) | 70 (60-86) |
| Radiologist | 100 (85-100) | 30 (13-53) | 59 (52-65) | 100 (85-100) | 65 (50-79) |

Abbreviations: PPV: Positive predictive value; NPV: Negative predictive value

**Table 2.** The proportion of false negative and false positive cases for DCNN model and radiologist (Test dataset N = 47).

|  | False negative (%) | False positive (%) |
| --- | --- | --- |
| DCNN model | 8/23 (35%) | 4/24 (17%) |
| Radiologist | 0/23 (0%) | 17/24 (71%) |

The ensemble model achieved the highest area under the receiver operating characteristic curve (AUC) of 0.76 (Figure 1). In comparison, the AUC of radiologist was 0.65. The decision threshold for the DCNN model was set at > 0.31 matching the best sensitivity maintaining reasonable specificity (Figure 2).

**Figure 1.**
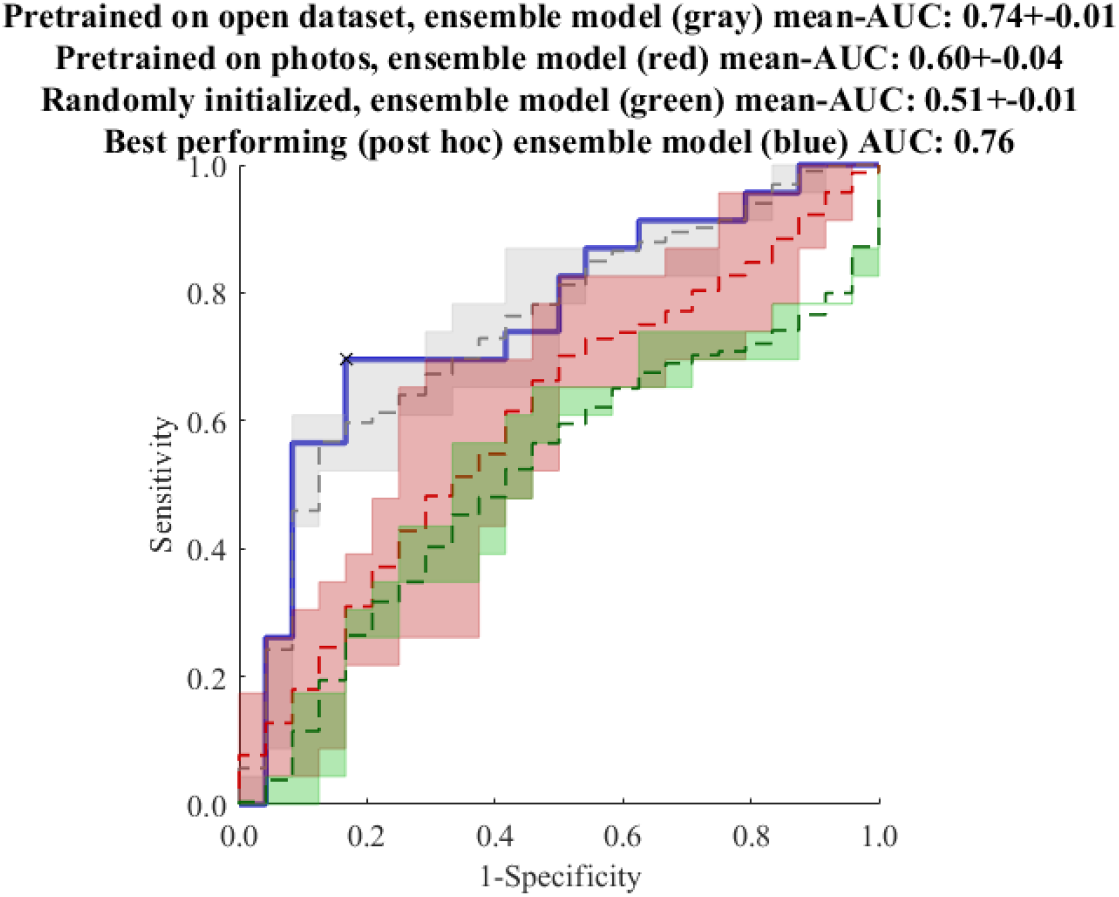
Receiver operating characteristic curves showing deep convolutional neural network (DCNN) model’s discrimination performance on the test dataset. The best performance of DCNN is achieved by ensemble model

**Figure 2.**
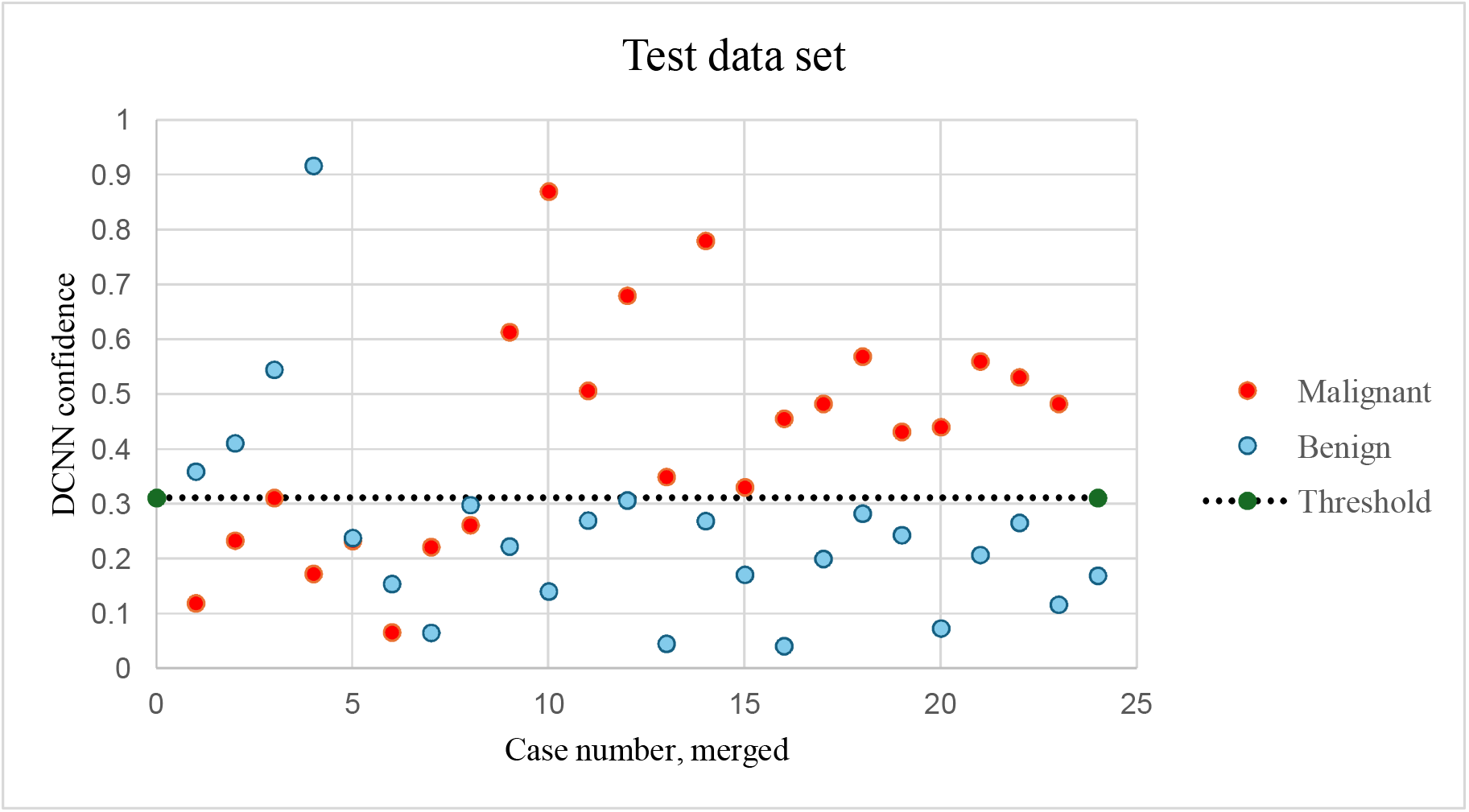
Deep convolutional neural network (DCNN) model output values (confidences) for the test dataset. The decision threshold (dashed line, output confidence ≥ 0.31 operating point) is optimal concerning sensitivity and specificity. Case numbers are merged to halve for better conspicuity.

Grad-CAM heatmaps revealed that, in all true positive cases identified by the DCNN model, predictions were based on the regions containing microcalcifications (Figure 3).

**Figure 3.**
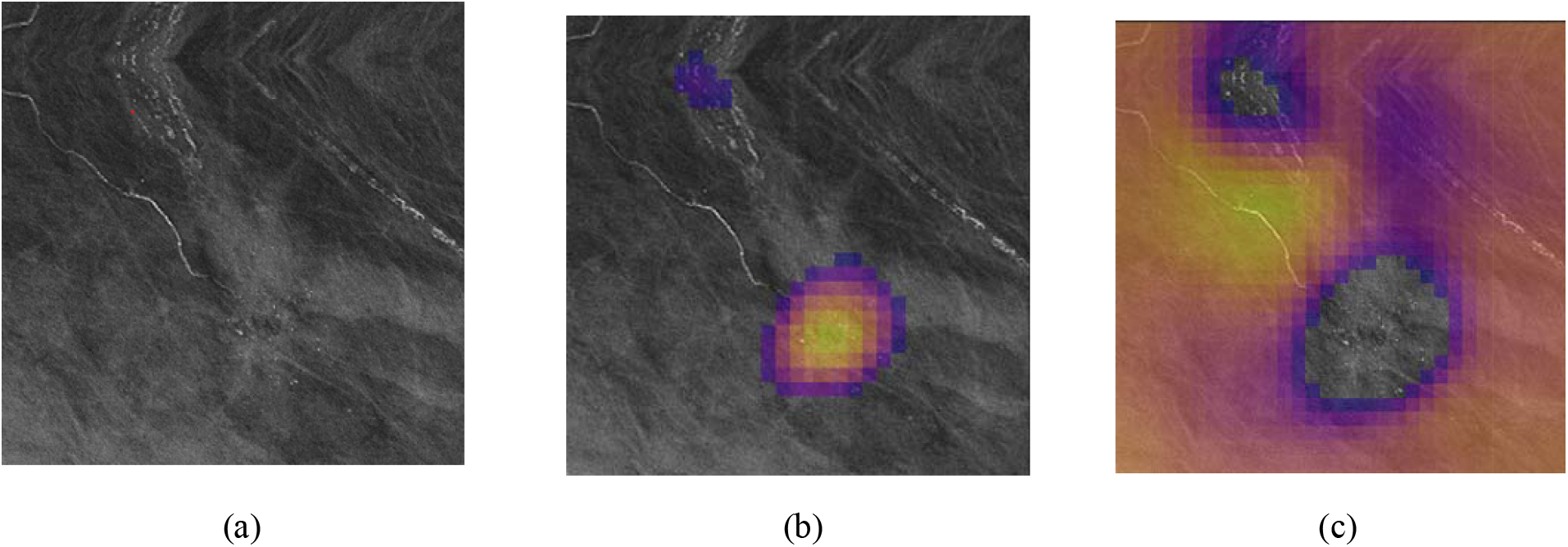
An example of Gradient-weighted Class Activation Mapping (Grad-CAM) heatmaps of true positive case with one of the highest probabilities for malignancy (0.78) given by DCNN model. Full-field digital mammography (FFDM) shows highly suspicious linear calcifications (a). Images (b) and (c) depict a heatmaps for malignant and benign activation respectively. Stereotactic vacuum-assisted core biopsy revealed ductal carcinoma in situ grade II.

False negatives included:

- Three cases with final pathology of risk lesion (classic lobular carcinoma in situ, atypical ductal hyperplasia). Grad-CAM heatmaps for these cases demonstrated activation of the correct region, but with probabilities below the cut-off threshold.
- Two DCIS grade I with correct region activation but probability at the threshold (0.31) or slightly beyond it (0.26)
- One case with correct region activation but a low probability score (0.06) (Figure 4).
- Two cases (4% of all test cases) involving invasive carcinoma foci in the final surgical specimens. These also exhibited correct region activation but insufficient probabilities (0.23 and 0.22).

False negative cases were primarily due to insufficient conspicuity of microcalcifications, caused by poor image quality or dense breast tissue.

**Figure 4.**
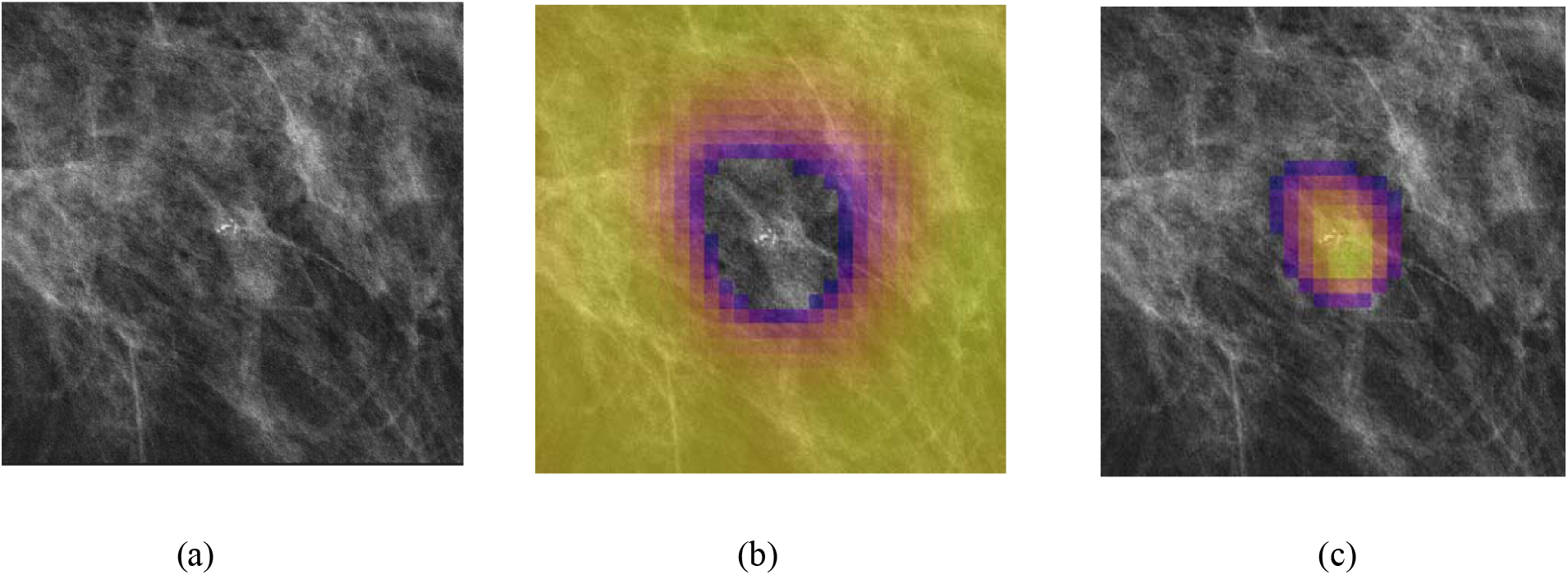
An example of Gradient-weighted Class Activation Mapping (Grad-CAM) heatmaps of false negative case with one of the lowest probabilities for malignancy (0.06) given by DCNN model. Full-field digital mammography (FFDM) shows (a) a group of fine pleomorphic calcifications (up to 38% rate of malignancy). Images (b) and (c) depict a heatmaps for malignant and benign activation respectively. Stereotactic vacuum-assisted core biopsy revealed ductal carcinoma in situ grade II.

False positives:

- Grad-CAM heatmaps for all false positive cases demonstrated activation of the correct region
- Two cases were classified as moderate suspicion (BIRADS 4b) by radiologist
- Two cases were classified as high suspicion (BIRADS 4c) by radiologist

## Discussion

We developed a novel DCNN algorithm, that utilizes d-TL approach and tested it for the characterization of suspicious microcalcifications found on mammograms. The ensemble model achieved an AUC of 0.76, with specificity and PPV values of 83% and 79%, respectively. Previous studies reported substantial inter-reader variability and low PPV for suspicious microcalcifications leading to a high rate of unnecessary biopsies (2,3). The results of the current study suggest that the DCNN algorithm may assist radiologists in determining the malignancy or benignity of microcalcifications, potentially reducing the need for additional imaging and unnecessary biopsies.

While the model’s sensitivity was moderate; the study focused on assessing the performance on already detected microcalcifications. In clinical practice, the DCNN model could serve as a decision-support tool (“intelligent spot” or i-spot) to assist radiologists in deciding whether to recall a patient for additional imaging or to proceed with biopsy. The optimal threshold was initially determined mathematically to maximize the AUC. However, adjusting the threshold, for example, downward to 0.23 could lead to the detection of three additional DCIS cases, including two with invasive components confirmed on final histopathology. This adjustment left two cases of ADH, one case of LCIS, and one outlier with a small invasive focus undetected. Lowering the threshold would increase the model’s sensitivity from 65% to 82%, but at the cost of reduced specificity (from 83% to 46%) and decreased overall accuracy (from 70% to 66%). The drop in specificity would result in 9 additional unnecessary biopsies.

Effective medical image analysis using DL methods requires access to large volumes of high-quality annotated training data. The need often conflicts with privacy and ethical concerns (20). TL offers a solution by leveraging pre-trained models to improve performance on target tasks with limited data available (21). However, even with TL, overfitting remains a concern when the model is fine-tuned on small datasets (22). The d-TL concept addresses the limitation by leveraging the similarity of source and target domains. Alkhaleefah et al. introduced a double-shot transfer learning technique, which demonstrated a significant improvement in the classification performance of a pre-trained network when tested on a public mammography dataset (23). Samala et al. showed that additional pre-training of DCNN model with mammograms improved the AUC over training with the DBT alone (so called multi-stage transfer learning) (24). To our best knowledge, this is the first study to apply the d-TL concept specifically to complex real-world clinical microcalcification cases. We included mammograms from different vendors to evaluate the model’s generalizability and minimize overfitting. The overall specificity and PPV of the DCNN model were significantly higher than those achieved using the BI-RADS lexicon.

XAI techniques, such as Grad-CAM, have gained attention for providing justifications, explanations, and insights into DL model predictions (25). In this study, Grad-CAM heatmaps confirmed that the DCNN model successfully identified regions containing suspected microcalcifications in all cases, despite image variability.

The study has several limitations. First, its single-centre retrospective study design introduces potential bias, as a DCNN model trained in a monocentric setting may not perform well when applied to different institutions with different patient demographics, disease prevalence, and clinical practices. Nonetheless, our dataset included diverse cases. Second, the training dataset was relatively small. While we utilized the d-TL technique to partially address this issue, the bias still exists, as the model may not generalize well to broader population. Future research with larger, more diverse datasets is needed to validate the effectiveness of the d-TL approach in broader population.

In conclusion, we demonstrated that a fine-tuned DCNN using d-TL achieved performance comparable to radiologists, with notably higher specificity and PPV, in classifying breast microcalcifications. The d-TL approach effectively addressed the limitations of small annotated datasets and integrating XAI enabled transparent evaluation of the model’s decision.

## Data Availability

All data produced in the present study are available upon reasonable request to the authors

